# The impact of vital signs on the death of patients with new coronavirus pneumonia: A systematic review and meta-analysis

**DOI:** 10.1101/2020.09.17.20196709

**Authors:** Meixia Du, Jie Zhao, Xiaochun Yin, Nadi Zhang, Guisen Zheng

**Author notes:** **Corresponding authors: Xiao-Chun Yin,PhD**, Gansu university of Chinese medicine, School of public health, Gansu university of Chinese medicine, Gansu Lanzhou, China., **Gui-Sen Zheng**, Gansu university of Chinese medicine, School of public health, Gansu university of Chinese medicine, Gansu Lanzhou, China.

## Abstract

**Background:** Assessing the impact of vital signs (blood pressure, body temperature, heart rate, respiratory rate, and oxygen saturation) on the death of patients with new coronavirus pneumonia would provide a simple and convenient method for the monitoring of subsequent illness, and therefore, in some degree reduce treatment costs and increase the cure rate clinically.

**Methods:** Six databases were retrieved. The software R 3.6.2 was used for meta-analysis of the included literature.

**Results:** 12 studies were included, which comprise 8996 patients affected with COVID-19 infection. The meta-analysis study found that blood pressure (MAP, SBP and DBP), heart rate, respiration rate and SpO2 are the risk factors for disease progression in patients with COVID-19. Among them, the increase in MAP and the decrease in SpO2 have the greatest impact on the death of patients with COVID-19 [MAP: MD = 5.66, 95% CI (0.34, 10.98), SpO2: MD = −5.87, 95% CI (−9.17, −2.57), P = 0.0005]. However, comparing the body temperature of the death group and the survival group found that the body temperature was not statistically significant between the two groups [body temperature: MD = 0.21, 95% CI (−0.01, 0.43), P = 0.0661].

**Conclusion:** The increase in MAP, heart rate and respiratory rate, as well as the decrease in SBP, DBP and SpO2 are all independent risk factors for death in patients with COVID-19. These factors are simple and easy to monitor, and individualized treatment can be given to patients in time, reducing the mortality rate and improving treatment efficiency.

## Background

In December 2019, a case of novel coronavirus pneumonia cause was reported in Wuhan, China ^[1-2]^. With the strong ability of transmission, the virus affects a wide range and propagates rapidly to other provinces and countries ^[3-5]^. According to the report of Wang et al. ^[6]^, the number of new coronaviruses diagnosed and died has been higher than that of SARS and Middle East Respiratory Syndrome (MERS) in 2003 and 2013 respectively. As of July 19, a total of 14284831 cases had been confirmed in 215 countries and regions, accounting for overall 602239 death and a high mortality rate of 4.22%. Strong spreading ability and large number of deaths have caused significant harm to human health and social economy. Many risk factors have been determined to be responsible for the death of the COVID-19 patients ^[7-10]^; however, the exact cause of death in the clinical treatment remains to be elucidated. As far as the prevention and treatment of the disease that is concerned, it is currently necessary to determine the factors that concisely and quickly predict the death of patients.

Meta-analysis, reviewing the published literature data, is a method to determine the risk factors in this paper. Meta-analysis execute a secondary analysis to expand the sample size, minimize the limitations of a single small sample clinical trial, to generate more comprehensive and reliable analysis results, thereby facilitating a better platform for medical decision-making. Therefore, it is an indispensable statistical method to determine the factors affecting the death of patients with COVID-19. Some literatures only analyze the impact of a certain disease or lifestyle on the death of patients with COVID-19. For example, Zhang et al. ^[11]^ studied the impact of the prevalence of hypertension on the death of patients with COVID-19, Vardavas et al. s^[12]^ analyzed the relationship between smoking and the adverse outcome of COVID-19. Vrsalovic et al. ^[13]^ used troponin to predict the death of COVID-19 patients. There are also some literatures that comprehensively analyzed multiple influencing factors, but there were only Chinese patients in these literatures, which caused a certain degree of one-sidedness. For example, Zheng et al. ^[14]^ conducted a comprehensive analysis of 13 Chinese literatures. analysis. And most of the current research results are about the basic demographic characteristics of patients and potential diseases. These factors could not be changed in generally, so that the symptoms could not be improved in the clinical cure. Although laboratory test results can predict the patient’s condition, they have certain delay, and it takes a long time and costs higher.

Analyzing comprehensively for most of the risk factors (vital signs) that affect the prognosis of COVID-19 patients, could reduce the mortality and give a reference for the surveillance of the of COVID-19 patients. So, in this paper, as many foreign patients as possible were included, which avoided the paper’s one-sideness, and the risk factors about vital signs (blood pressure, body temperature, heart rate, respiratory rate, and oxygen saturation), which were simpler and more convenient then experimental tests in the lab, were analyzed.

## Methods

### Search policy and selection criteria

Applying the method of combining the keywords and the abstracts, six databases including PubMed, Scopus, Embase, CNKI, China biomedical literature database (CBM), and WanFang data knowledge service platform were explored by computer to assemble relevant studies analyzing the factors affecting the death of COVID-19 published from January 1, 2020, to July 16, 2020. were also reviewed to obtain relevant studies. In this search process, there was no language limitation. Terms included: 2019-nCoV or Wuhan coronavirus or SARS-CoV-2 or 2019 novel coronavirus or COVID-19 virus or coronavirus disease 2019 virus or Wuhan seafood market pneumonia virus and death or Mortalities or Mortality or Fatality and blood pressure or body temperature or heart rate or respiratory rate (eg: ((((((((2019-nCoV [Title/Abstract]) OR (Wuhan coronavirus[Title/Abstract])) OR (SARS-CoV-2[Title/ Abstract])) OR (2019 novel coronavirus[Title/Abstract])) OR (COVID-19 virus[Title/ Abstract])) OR (coronavirus disease 2019 virus[Title/Abstract])) OR (Wuhan seafood market pneumonia virus[Title/Abstract])) AND (((death[Title/Abstract]) OR (mortality[Title/Abstract])) OR (fatality[Title/Abstract]))) AND ((((blood pressure [Title/Abstract]) OR (respiratory rate[Title/Abstract])) OR (heart rate[Title/Abstract])) OR (body temperature [Title/Abstract])).(The detailed search strategies from databases).

The inclusion criteria were as follows: (1) COVID-19 diagnosed patients; (2) the endpoint was well-defined, involving death and non-death patients; (3) study designs included randomized controlled trials, nonunionized controlled trials, case-control studies, cohort studies, cross-sectional studies, and also case reports; (4) the sample size of the study was greater than 20.

The exclusion criteria involved were as follows: (1) republished literature; (2) literature irrelevant to the research purpose; (3) literature review; (4) no control group; (5) literature in which valid data were not obtained or data could not be used.

### Data Extraction

Two researchers (Du MX & Zhang ND) independently completed literature selection, data extraction, and cross-checking using Endnote 8.0 and Microsoft Excel software. Another researcher (Yin XC & Zheng GS) was responsible for resolving any sort of disagreement. The database was developed with the aid of Microsoft Excel and the data extracted primarily included: the first author, publication time, the study design type, vital signs, and the detailed information of markers.

### Quality Assessment

The MINORS ^[15]^ was applied to assess bias risk. There are 12 entries in this standard, each of which gets a score of 0 to 2, in which 0 represents unreported, 1 designates reported but insufficient information, and 2 signifies reported and provided sufficient information. The greater the total score of each item, the higher the quality of the literature. Among them, a score of ≥ 19 is classified as high quality, 13-18 as medium quality, and ≤ 12 as low quality. Studies with scores lower than 12 were excluded from the final meta-analysis. The process of literature evaluation was compiled by two researchers independently and in case of disagreement, by the third researcher.

### Statistical Analysis of Data

Statistical analysis of the data was carried out with the help of R 3.6.2 software. The results of the included studies were performed with fixed-effect models (Mantel–Haenszel method) or random effect models in the cases of significant heterogeneity between studies. We used *I*^*2*^ and Q tests to test the heterogeneity of the included literature. No statistical heterogeneity was considered if *p* > 0.10 or *I*^*2*^ < 50%, and the fixed-effect model, i.e., Mantel–Haenszel method (M-H method), was used for analysis. If 0.05 < *p* < 0.10 and *I*^*2*^ > 50%, or *p* ≤ 0.05, there was statistical heterogeneity. The source of heterogeneity was first analyzed, and a low-quality study with large bias was excluded, and the combined effect size was computed again. Even after the above treatment, if the results of multiple studies still displayed heterogeneity, the random effect model, namely, the Dersimonian-Laird method (D-L method), was applied for meta-analysis. In order to increase the number of studies, according to the Cochrane 4.2 user manual, when the sample size was large (n ≥ 30), the median was used as the mean value, SD = (upper-lower limit)/1.35, and the data from the included literature were analyzed using mean and SD. Test level, α = 0.05.

## Results

### Research Selection and Quality Assessment

Based on the search strategy, 251 studies were screened from the online database. A total of 175 records were retained after removing duplicate records. Thereafter, 119 articles were excluded by the titles and abstracts, and 43 of the remaining 56 articles were deleted for various reasons, and we were left with 13 articles. According to the literature quality evaluation, 12 articles of 3493 patients ^[16–28]^ were ultimately selected for the meta-analysis (Figure 1). All these selected studies were published in 2020 with varying sample sizes ranging from 20 to 7614 patients. The risk of bias and applicability concerns for the included studies are detailed in Table 1. According to the MINORS assessment, none of the studies was considered to be seriously flawed. The score of the 12 included studies ranged between 18 and 22. Thus all studies were considered to have a low risk of bias for selection.

**Table 1.** Main characteristics of the included studies.

| Study | Year | Research type | Country | Number of Patients(n) | Male(n) |  | Age(years) |  | Quality evaluation score (score) |
| --- | --- | --- | --- | --- | --- | --- | --- | --- | --- |
|  |  |  |  |  | Deaths | Survivors | Deaths | Survivors |  |
| Lang Wang<sup>[18]</sup> | 2020 | Retrospectives Study | China | 202 | 23 | 65 | 74.0(65.0, 84.0) | 61.0(49.0, 67.0) | 18 |
| Lang Wang<sup>[17]</sup> | 2020 | Retrospectives Study | China | 339 | 39 | 130 | 76.0 (70.0, 83.0) | 68.0 (64.0, 74.0) | 21 |
| Tao Chen<sup>[19]</sup> | 2020 | Retrospectives Study | China | 274 | 83 | 88 | 68.0(62.0, 77.0) | 51.0(37.0, 66.0) | 18 |
| Hai Hu<sup>[11]</sup> | 2020 | Retrospectives Study | China | 105 | 14 | 48 | 75.05 ± 12.94 | 57.71 ± 15.34 | 19 |
| Min Li | 2020 | Retrospectives Study | China | 46 | 10 | 26 | 68.2 ± 10.9 | 54.5 ± 15.5 | 18 |
| Jiayin Lu | 2020 | Retrospectives Study | China | 20 | 5 | 3 | 69.7 ± 15.6 | 69.8 ± 7.79 | 18 |
| Hang Yang | 2020 | Retrospectives Study | China | 94 | 8 | 37 | 77.0 ( 675, 83.0) | 66.0(59.0, 72.5) | 18 |
| Feng Pan<sup>[23]</sup> | 2020 | Retrospectives Study | China | 124 | 67 | 18 | 69.0 (61.0, 73.0) | 65.0(49.0, 77.0) | 19 |
| Serena Tharakan | 2020 | Retrospectives Study | America | 7614 | 765 | 3353 | 73.8 ± 12.5 | 56.5 ± 18.1 | 18 |
| Marcello • covino<sup>[24]</sup> | 2020 | Retrospectives Study | Italy | 69 | 12 | 25 | 85.0 (83.0, 86.0) | 84.0 (81.0, 89.0) | 19 |
| Qingchun Yao<sup>[27]</sup> | 2020 | Retrospectives Study | China | 25 | 7 | 6 | 65(51.0, 73.5) | 56(50.5, 63.5) | 18 |
| Chaomin Wu<sup>[15]</sup> | 2020 | Retrospectives Study | China | 84 | 29 | 31 | 68.5 (59.3, 75.0) | 50.0 (40.3, 56.8) | 22 |

**Figure 1.**
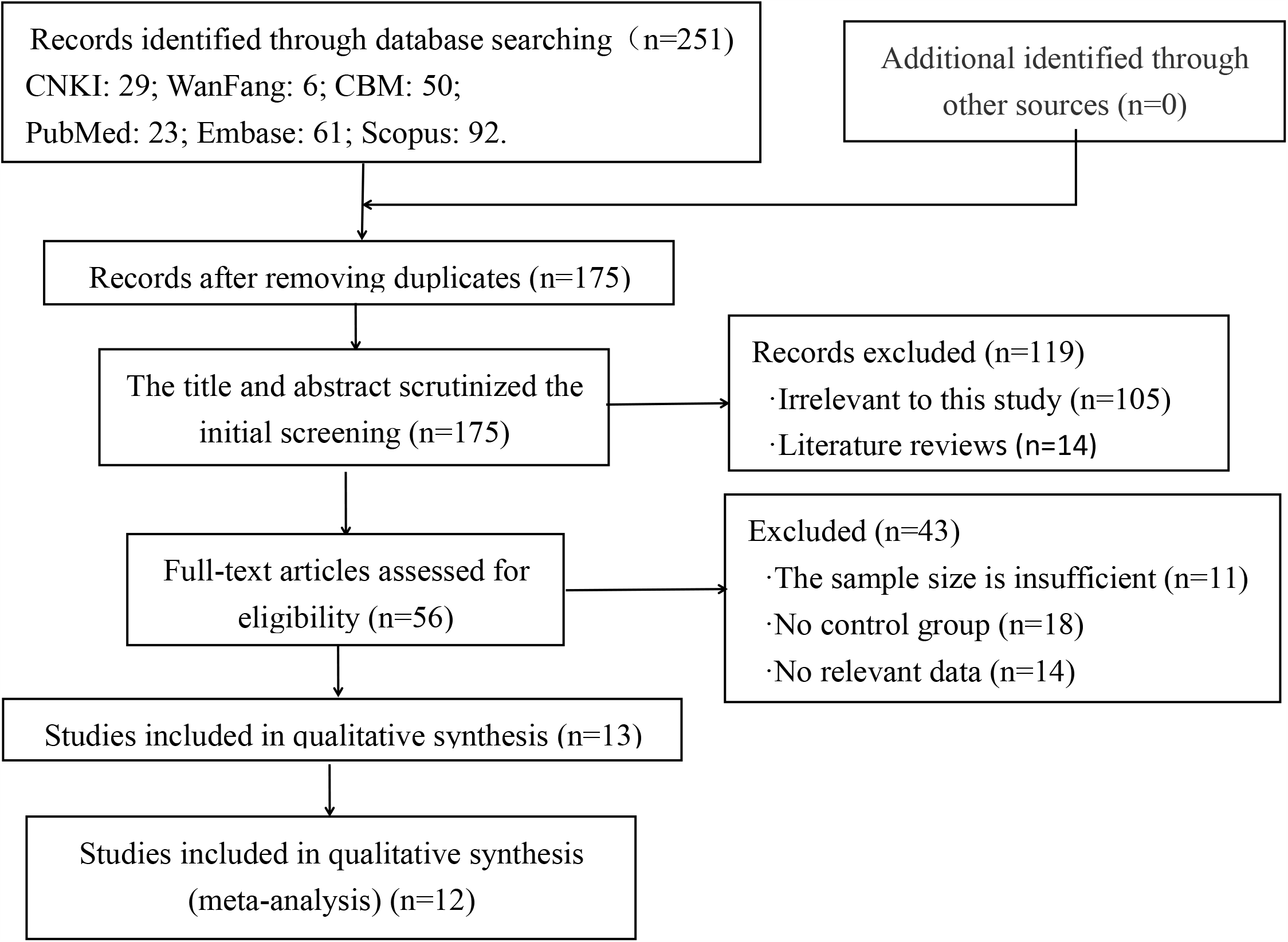
Flow chart of the literature screening process.

### Blood pressure

The demographic characteristics of the included literature were illustrated in Table 1 for meta-analysis. The median age ranged from 51.0 to 86.0 years for the patients in the death group, whereas 40.3 to 89.0 years for the survivor group. Heterogeneity test was performed initially, the relevant literature has no statistical heterogeneity in MAP(Mean artery pressure) and SBP(Systolic blood pressure), but there is statistical heterogeneity between DBP(Diastolic blood pressure)-related literature (MAP: *p* = 0.80, *I*^*2*^ = 0%; SBP: *p* = 0.10, *I*^*2*^ = 49%; DBP: *p* = 0.03, *I*^*2*^ = 58%;). Therefore, the fixed-effect model is used for meta-analysis of MAP and SBP, while the analysis of DBP uses the random effects model. The results showed that the MAP value of the death group was significantly higher than that of the survival group, while SBP and DBP were significantly lower than the survival group, and the differences were statistically significant [MAP: MD = 5.66, 95% CI (0.34, 10.98), P = 0.0371; SBP: MD = −2.30, 95% CI (−3.80, −0.80), P = 0.0027; DBP: MD = −3.00, 95% CI (−5.51, −0.48), P = 0.0194] (Figure 2).

**Figure 2.**
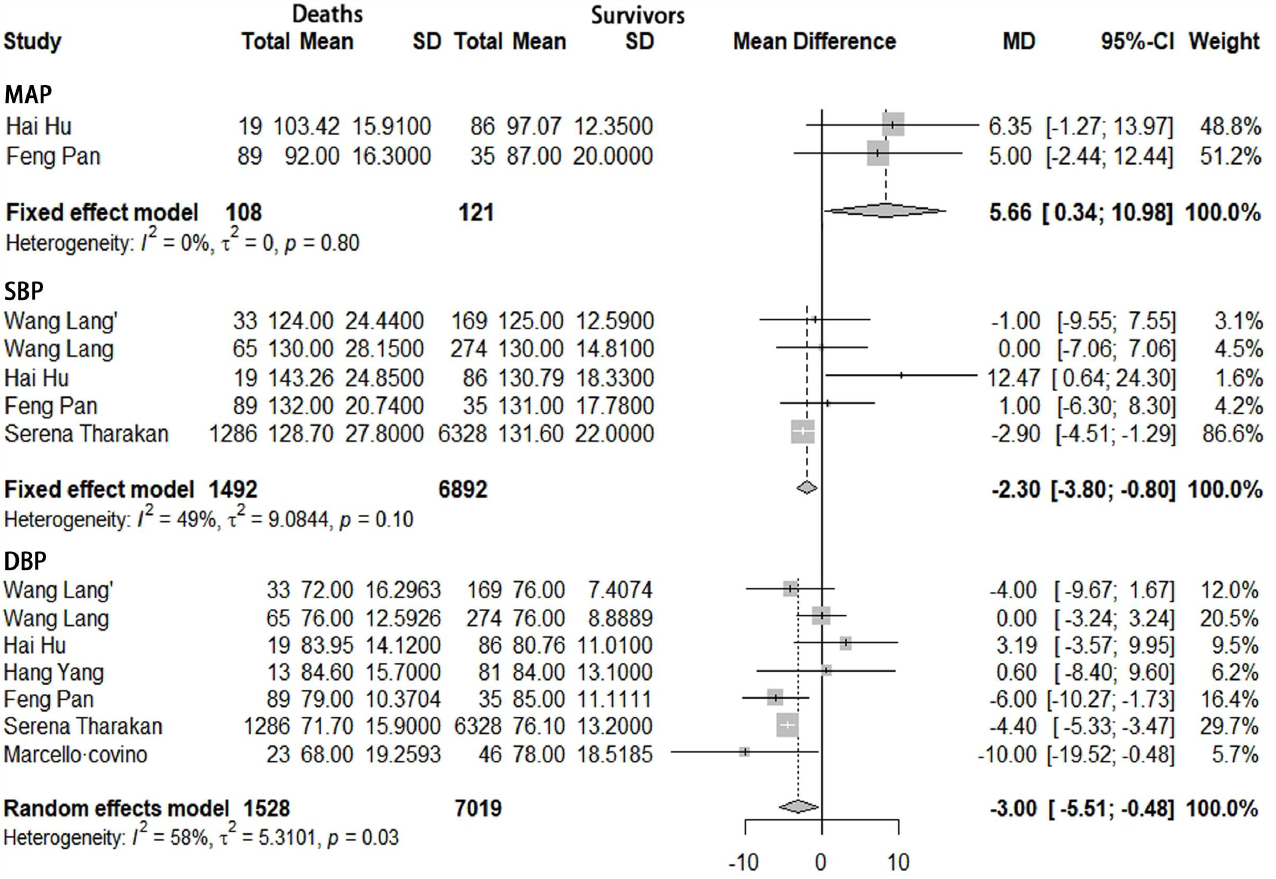
Meta-analysis for MAP, SBP and DBP in COVID-19 cases. Heterogeneity analysis was carried out using Q test for the variation among the studies (I2 index). Forest plots depicted the comparison of the incidence of MAP, SBP and DBP in death group and survivor group.

### Body temperature

The prevalence of body temperature included in the literature was considered in the meta-analysis to elucidate their impact on the death of COVID-19. Estimation of the heterogeneity of the selected articles showed that there was not any statistical heterogeneity among the literature related to body temperature (*p =* 0.74, *I*^*2*^ *=* 0%). Thus, for further investigation, the fixed-effect model was implemented. Statistical analysis showed that the body temperature of the death group was higher than that of the survival group, but the difference between the two groups was not statistically significant [body temperature: MD = 0.21, 95% CI (−0.01, 0.43), P = 0.0661] (Figure 3).

**Figure 3.**
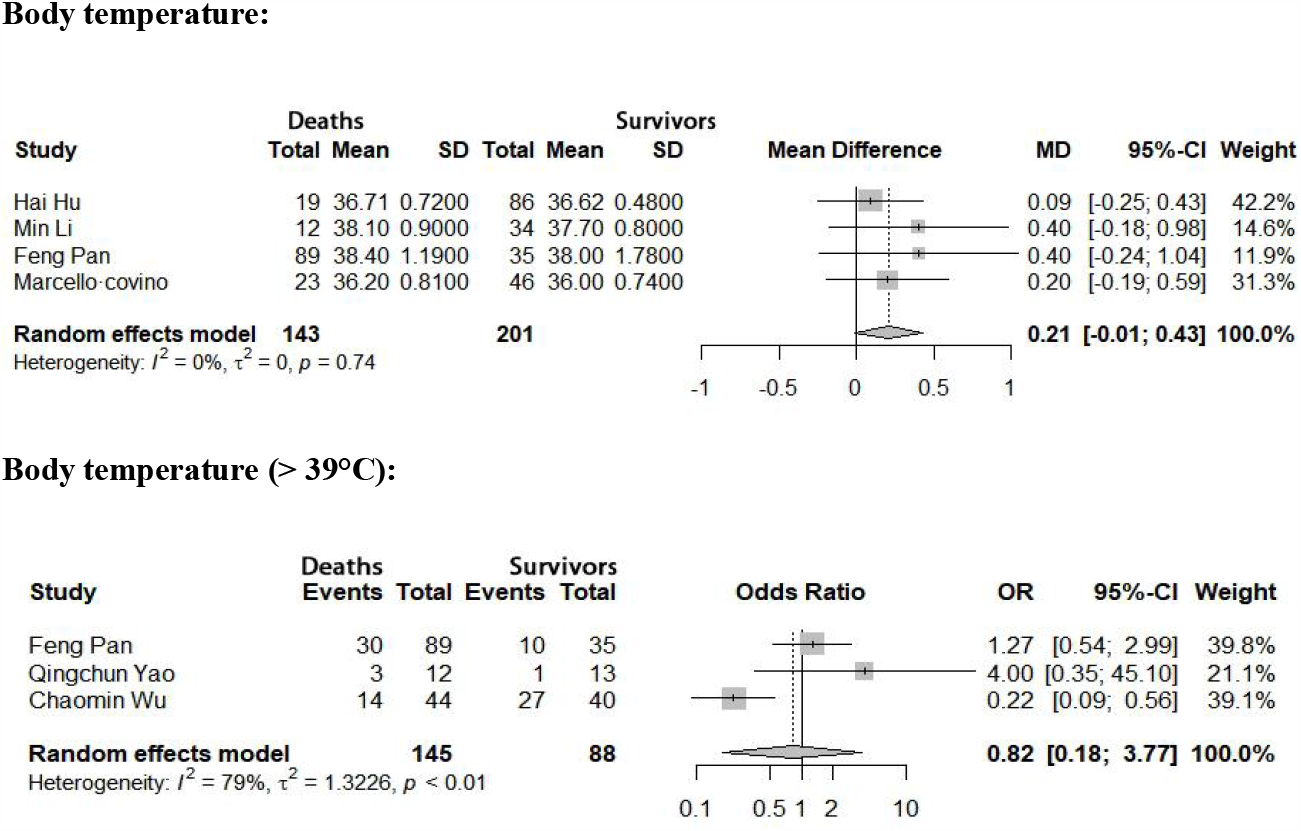
Meta-analysis for body temperature in COVID-19 cases. Heterogeneity analysis was carried out using Q test for the variation among the studies (I2 index). Forest plots depicted the comparison of the incidences of body temperature in the death group and survivor group.

In order to further analyze whether body temperature has an impact on the death of patients with new coronavirus pneumonia, this article conducted a meta-analysis of the proportion of the dead group and the survival group with a body temperature of > 39°C. First, the heterogeneity test found that there are heterogeneous differences between the relevant documents (*p <* 0.701, *I*^*2*^ *=* 79%), so the random effects model is used for analysis. The analysis results showed that the proportion of patients with body temperature (> 39°C) in the death group was lower than that in the survival group, but the difference between the two groups was not statistically significant [body temperature (> 39°C): OR = 0.82, 95% CI (0.18, 3,77), P = 0.8017] (Figure 3).

### Heart rate

To determine the effect of the heart rate as a risk factor for death of patients suffering from COVID-19, a meta-analysis was performed on the vital signs of the deaths and the survivors of 8 included articles (with a total amount of 8544 patients). The fixed-effect model could be used for analysis as there was no statistical heterogeneity of heart rate among the pertinent literature (*p* = 0.11, *I*^*2*^ = 40%) based on the heterogeneity test. The heart rate of the death group was considered to be significantly higher than that of the survivor group [heart rate: MD = 2.33, 95% CI (1.20, 3.46), P < 0.0001] (Figure 4).

**Figure 4.**
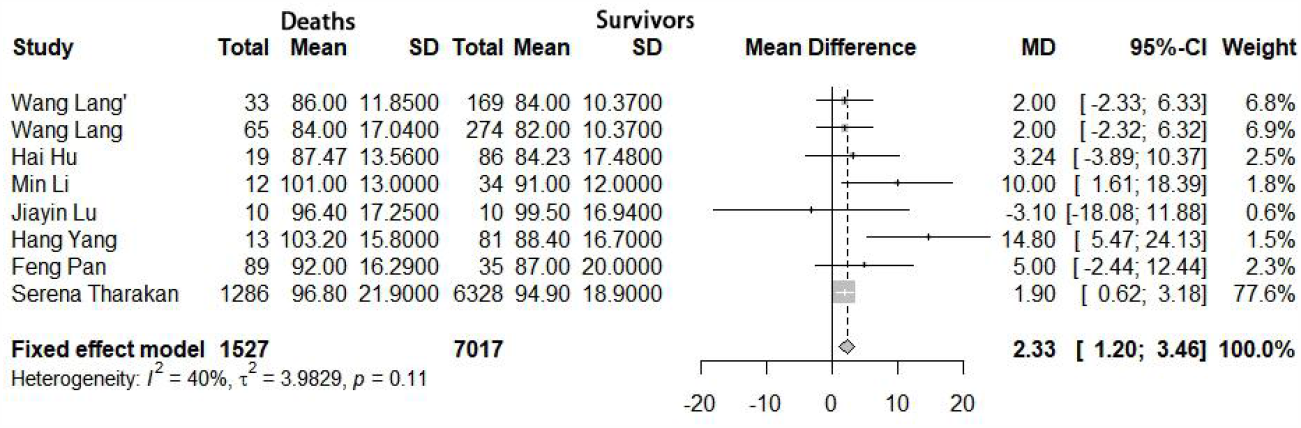
Meta-analysis for heart rate in COVID-19 cases. Heterogeneity analysis was carried out using Q test for the variation among the studies (I2 index). Forest plots depicted the comparison of the incidences of heart rate in the death group and survivor group.

### Respiratory rate

Meta-analysis was also executed on the respiratory rate manifested by the two groups of COVID-19 patients-death and survivor-in the included 6 literature. Statistically significant heterogeneity among the literature related to respiratory rate (*p* = 0.11, *I*^*2*^ = 45%) was absent as per the heterogeneity test, so the fixed-effect model was applied for meta-analysis. Statistical evaluation verified that the respiratory rate of the death group was significantly higher than that of the survival group, and there was statistical significance [respiratory rate: MD = 2.66, 95% CI (1.49, 3.83), P = 0.0048] (Figure 5).

**Figure 5.**
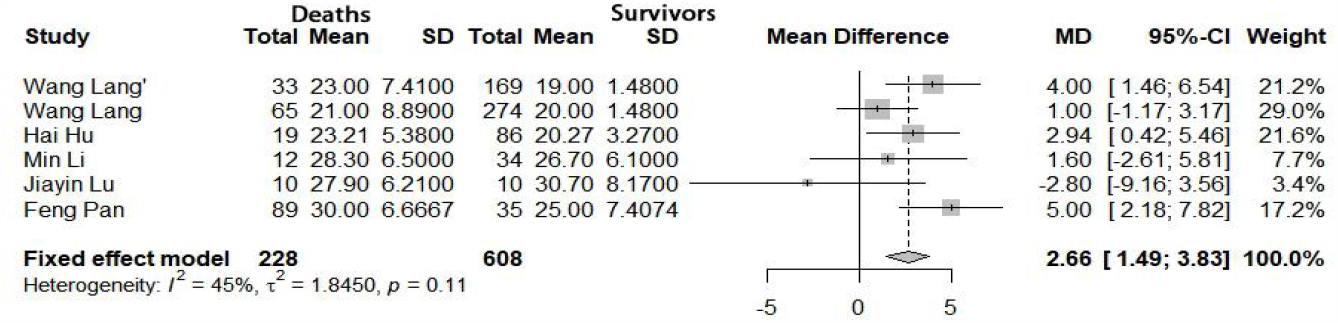
Meta-analysis for respiratory rate in COVID-19 cases. Heterogeneity analysis was carried out using Q test for the variation among the studies (I2 index). Forest plots depicted the comparison of the incidences of respiratory rate in the death group and survivor group.

### Oxygen saturation (SpO2)

Analyze the difference in SpO2 between the death group and the survival group in the 6 included literatures to determine the impact of blood oxygen saturation on the death of patients with COVID-19 Heterogeneity inspection results to SpO2 (*P* < 0.01, *I*^*2*^ = 91%) estimated there were heterogeneity differences among related to literature, so the random effects model was adopted for meta-analysis. The outcomes of statistical analysis witnessed the SpO2 of the death group were significantly lower than that of the survivor group [SpO2: MD = −5.87, 95% CI (−9.17, −2.57), P = 0.0005] (Figure 6).

**Figure 6.**
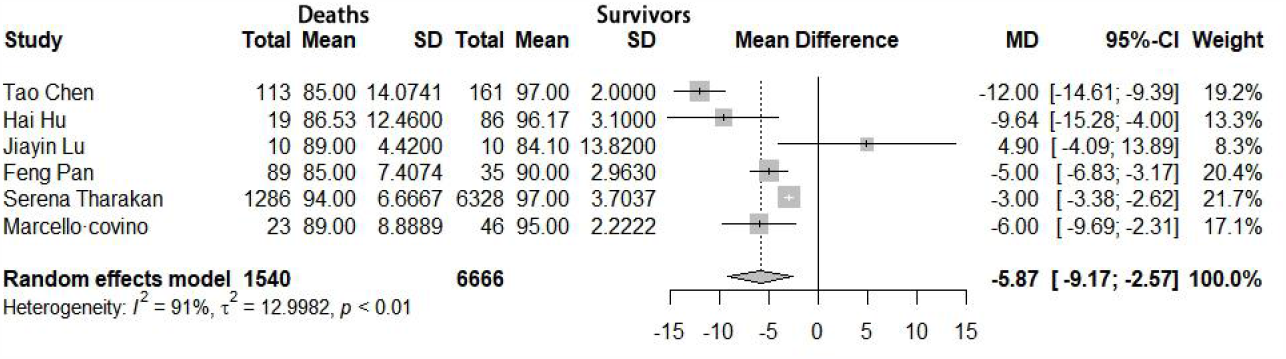
Meta-analysis for the SpO2 in COVID-19 cases. Heterogeneity analysis was carried out using Q test for the variation among the studies (I2 index). Forest plots depicted the comparison of the incidence of SpO2 in the death group and survivor group.

## Discussion

Novel coronavirus disease (COVID-19) is another serious threat to public health after SARS, Ebola, and avian flu. Owing to its fast transmissibility through respiratory droplets and close contact, the virus is extremely infectious and pathogenic ^[28-29]^, thereby making COVID-19 a global concern within a short period. This article mainly conducts a meta-analysis of the impact of vital signs on the death of patients with COVID-19, and evaluates the impact of vital signs on the prognosis of COVID-19. To facilitate the monitoring of changes in the condition of COVID-19 patients, provide personalized treatment, reduce the mortality rate, and increase treatment benefits.

This study reported that the MAP of deaths from COVID-19 was higher than that of survivors, while SBP and DBP were lower than those of survivors. The results showed that the MD values of MAP, SDP, and DBP of the death group were 5.66, −2.30, −3.00, and all of them were statistically significant. This shows that the increase in MAP and the decrease in SDP and DBP will increase the risk of death from COVID-19. According to previous studies ^[30-32]^, it has been found that patients with COVID-19 will have complications such as shock and acute myocardial injury, and the proportion of complications in the death group is higher than that in the survival group, and these will cause the patient’s systolic and arterial pressure to drop. Blood pressure measurement is relatively simple and convenient, and detecting changes in blood pressure can give patients personalized treatment early and reduce the mortality rate.

Body temperature is one of the variables included in the pneumonia severity index ^[33]^ and systemic inflammatory response syndrome (SIRS) ^[34]^. In this study, a meta-analysis of the body temperature of the death group and the survival group included in 4 literatures showed that the body temperature of the death group was higher than that of the death group, but the difference between the two was not statistically significant. Fever is the most common symptoms of patients with COVID-19 infection and the research factor with the highest incidence of initial symptoms in admitted patients. Therefore, the difference in body temperature between the two groups was not significant. Fever is a general clinical feature of patients with COVID-19 on admission. Therefore, most patients will have symptoms of fever and have different body temperatures. In order to further study whether body temperature will affect the prognosis of patients with COVID-19, this article discusses the death group and The proportion of body temperature > 39°C in the survival group was analyzed, and it was found that the proportion of body temperature > 39°C in the death group was lower than that in the survival group, but the difference between the two groups was not statistically significant. Some studies found that low body temperature at the initial presentation is a marker of poor prognosis, and high maximum temperature during the course of SARS-CoV-2 infection was a significant harbinger of poor outcomes ^[24]^. Therefore, body temperature has a great influence on the prognosis of patients with COVID-19. The patient’s body temperature should be monitored regularly at an early stage, and personalized treatment should be given early to improve treatment benefits.

The heart rate of the patients was examined, and the MD value of the heart rate of the deceased was equal to 2.33 compared with that of the survivor group. This indicated increased death risk of the patients with accelerated heart. Accelerated heart rate indicates the damage of myocardial energy supply, and one of the complications of patients with COVID-19 is acute myocardial injury. Study has shown that in addition to acute respiratory distress syndrome (ARDS), systemic inflammation with cardiac dysfunction is the main cause of death from severe COVID-19 ^[23]^. So early detection of changes in heart rate and prevention of complications can decrease the mortality of the disease.

This study found that the respiratory rate of the COVID-19 death group was greater than of the survival group, and SpO2 was lower than that of the survival group. Compared with the survival group, the MD values of respiratory rate and SpO2 in the death group were 2.66 and −5.87, respectively. An increase in respiratory rate and a decrease in SpO2 indicates lung damage.Studies have found that the Sars-Cov2 cell entry receptor is located mainly in the lungs, more than half of patients might develop dyspnea, and >10% might require ventilatory support ^[35-37]^. With acute hypoxia being the main determinant of disease progression and severity, the SpO2 of admission is crucial for death risk stratifification ^[25]^. Therefore, an increase in respiratory rate and a decrease in SpO2 will indicate lung function damage, worsening of the disease, and death. Monitoring methods of respiratory rate and SpO2 are simple and convenient. And can be used as predictors of poor prognosis for patients with COVID-19. Improve clinical symptoms as soon as possible, prevent complications, and improve treatment benefits.

## Conclusion

In this study, a meta-analysis was conducted to study the impact of the vital signs of patients with COVID-19 on their death. The results found that the increase in MAP, heart rate and respiratory rate, as well as the decrease in SBP, DBP and SpO2 are all independent risk factors for death in patients with COVID-19. Monitoring methods for these factors are simple and convenient, which can reduce medical costs and increase treatment benefits. Thus individualized treatment should be given in time and attention should be paid to the risk factors, to improve the efficacy and reduce the mortality.

## List of abbreviations

COVID-19: Novel coronavirus; MERS: Middle East respiratory syndrome; MAP: Mean artery pressure; SBP: Systolic blood pressure; DBP: Diastolic blood pressure; SpO2: Oxygen saturation

## Data Availability

The data of COVID-19 are available from published literature. All data used for analysis are available upon a proper request from the corresponding author Xiaochun Yin at.

## Declarations

## Acknowledgements

The authors thank all medical staff and health practitioners who have contributed to the reporting of human COVID-19.

## Authors’ contributions

MXD and XCY contributed to the conception and design of the study, had full access to all data in the study and take responsibility for the integrity and accuracy of data analysis. MXD and NDZ contributed to data acquisition. MXD, NDZ, XCY and GSZ contributed to the analysis and interpretation of the data. All authors participated in manuscript writing and revision and approved the final version of the manuscript.

## Availability of data and materials

The data of COVID-19 are available from published literature. All data used for analysis are available upon a proper request from the corresponding author Xiaochun Yin a<u>t</u>.

## Ethics approval and consent to participate

Not applicable.

## Consent for publication

Not applicable.

## References

[1] Tang JW, Tambyah PA, Hui DSC. Emergence of a novel coronavirus causing respiratory illness from Wuhan, China. J Infect [Internet]. 2020;80(3):350–371. doi:10.1016/j.jinf.2020.01.014.

[2] Zhou P, Yang XL, Wang XG, et al. A pneumonia outbreak associated with a new coronavirus of probable bat origin. Nature [Internet]. 2020;579(7798):270–273. doi: 10.1038/s41586-020-2012-7.

[3] Carlos WG, Dela Cruz CS, Cao B, et al. Novel Wuhan (2019-nCoV) Coronavirus. Am J Respir Crit Care Med [Internet]. 2020;201(4):P7–P8. doi:10.1164/rccm.2014P7.

[4] Lu H. Drug treatment options for the 2019-new coronavirus (2019-nCoV). Biosci Trends [Internet]. 2020;14(1):69–71. doi:10.5582/bst.2020.01020.

[5] Yan Y, Shin WI, Pang YX, et al. The First 75 Days of Novel Coronavirus (SARS-CoV-2) Outbreak: Recent Advances, Prevention, and Treatment. Int J Environ Res Public Health [Internet]. 2020;17(7):2323. Published 2020 Mar 30. doi:10.3390/ijerph17072323.

[6] Wang Y, Wang Y, Chen Y, et al. Unique epidemiological and clinical features of the emerging 2019 novel coronavirus pneumonia (COVID-19) implicate special control measures. J Med Virol [Internet]. 2020;10.1002/jmv.25748. doi:10.1002/jmv.25748.

[7] Du RH, Liang LR, Yang CQ, et al. Predictors of mortality for patients with COVID-19 pneumonia caused by SARS-CoV-2: a prospective cohort study. Eur Respir J [Internet]. 2020;55(5):2000524. Published 2020 May 7. doi:10.1183/13993003.00524-2020.

[8] [An W, Xia F, Chen M, et al. Clinical characteristics of 11 patients with COVID-19 death. Journal of practical medicine [Internet]: 1-6[2020-05-07]. http://kns.cnki.net/kcms/detail/44.1193.r.20200414.1620.007.html.]

[9] Li Y, Yang Z, Ai T, et al. Association of “initial CT” findings with mortality in older patients with coronavirus disease 2019 (COVID-19). Eur Radiol [Internet]. 2020;1–8. doi:10.1007/s00330-020-06969-5.

[10] Paranjpe I, Russak A, De Freitas JK, et al. Clinical Characteristics of Hospitalized Covid-19 Patients in New York City. Preprint. medRxiv [Internet]. 2020;2020. 04.19.20062117. Published 2020 Apr 23. doi:10.1101/2020.04.19.20062117.

[11] Zhang J, Wu J, Sun X, et al. Association of hypertension with the severity and fatality of SARS-CoV-2 infection: A meta-analysis. Epidemiol Infect [Internet]. 2020;148:e106.Published 2020 May 28. doi:10.1017/S095026882000117X.

[12] Vardavas CI, Nikitara K. COVID-19 and smoking: A systematic review of the evidence. Tob Induc Dis. 2020;18:20. Published 2020 Mar 20. doi:10.18332/tid/119324.

[13] Vrsalovic M, Vrsalovic Presecki A. Cardiac troponins predict mortality in patients with COVID-19: A meta-analysis of adjusted risk estimates. J Infect [Internet]. 2020;S0163-4453(20) 30300-5. doi:10.1016/j.jinf.2020.05.022.

[14] Zheng Z, Peng F, Xu B, et al. Risk factors of critical & mortal COVID-19 cases: A systematic literature review and meta-analysis [published online ahead of print, 2020 Apr 23.

[15] Slim K, Nini E, Forestier D, Kwiatkowski F, et al. Methodological index for non-randomized studies (minors): development and validation of a new instrument. ANZ J Surg [Internet]. 2003;73(9):712–716.doi:10.1046/j.1445-2197.2003.02748.x.

[16] [Lang W, Wen H, Yu X, et al. Effects of myocardial injury on clinical prognosis of COVID-19 patients. Chinese journal of cardiovascular disease [Internet], 2020, 48 (2020-04-14). http://rs.yiigle.com/yufabiao/1188751.htm.DOI: 10.3760/cma.j.cn112148-20200313-00202.]

[17] Wang L, He W, Yu X, et al. Coronavirus disease 2019 in elderly patients: Characteristics and prognostic factors based on 4-week follow-up. J Infect [Internet]. 2020; 80(6): 639–645. doi: 10.1016/j.jinf.2020.03.019.

[18] Chen T, Wu D, Chen H, et al. Clinical characteristics of 113 deceased patients with coronavirus disease 2019: retrospective study. BMJ [Internet]. 2020;368:m1091. Published 2020 Mar 26. doi:10.1136/bmj.m1091.

[19] Hu H, Yao N, Qiu Y. Comparing Rapid Scoring Systems in Mortality Prediction of Critically Ill Patients With Novel Coronavirus Disease. Acad Emerg Med [Internet]. 2020; 10.1111/ acem. 13992. doi:10.1111/acem.13992.

[20] Li M, Liu JX, Hu QB. Clinical characteristics of patients with severe COVID-19 and influencing factors of death. Journal of Practical Cardio-Cerebral and Pulmonary Vascular Diseases, 20,28(06):1–5.

[21] Lu JY, Zhang Y, Cheng G, et al. Clinical characteristics and outcome analysis of adult covid-19 patients in ICU in honghu, hubei province. Journal of southern medical university,2020,40(06):778–785.

[22] Yang H, Yang LC, Zhang RT, et al. Beijing Da Xue Xue Bao Yi Xue Ban. 2020;52(3):420–424. doi:10.19723/j.issn.1671-167X.2020.03.004.

[23] Pan F, Yang L, Li Y, et al. Factors associated with death outcome in patients with severe coronavirus disease-19 (COVID-19): a case-control study. Int J Med Sci. 2020;17(9):1281–1292. Published 2020 May 18. doi:10.7150/ijms.46614.

[24] Tharakan S, Nomoto K, Miyashita S, et al. Body temperature correlates with mortality in COVID-19 patients. Crit Care. 2020;24(1):298. Published 2020 Jun 5. doi:10.1186/s13054-020-03045-8.

[25] Covino M, De Matteis G, Santoro M, et al. Clinical characteristics and prognostic factors in COVID-19 patients aged ≥80?years. Geriatr Gerontol Int. 2020;10.1111/ggi.13960.doi:10.1111/ggi.13960.

[26] Yao Q, Wang P, Wnag X, et al. Retrospective study of risk factors for severe SARS-Cov-2 infections in hospitalized adult patients. Pol Arch Intern Med [Internet]. 2020;130(5):390–399. doi:10.20452/pamw.15312.

[27] Wu C, Chen X, Cai Y, et al. Risk Factors Associated With Acute Respiratory Distress Syndrome and Death in Patients With Coronavirus Disease 2019 Pneumonia in Wuhan, China. JAMA Intern Med [Internet]. 2020;e200994. doi:10.1001/jamainternmed.2020.0994.

[28] Kang M, Wu J, Ma W. Evidence and characteristics of human-to-human transmission of SARS-CoV-2. medRxiv [Internet]. 2020.https://doi.org/10.1101/2020.02.03.20019141.

[29] Riou J, Althaus CL. Pattern of early human-to-human transmission of Wuhan 2019 novel coronavirus (2019-nCoV), December 2019 to January 2020. Euro Surveill [Internet]. 2020;25(4):p 2000058. doi:10.2807/1560-7917.ES.2020.25.4.2000058.

[30] Cao J, Tu WJ, Cheng W, et al. Clinical Features and Short-term Outcomes of 102 Patients with Corona Virus Disease 2019 in Wuhan, China [published online ahead of print, 2020 Apr 2]. Clin Infect Dis. 2020;ciaa243. doi:10.1093/cid/ciaa243.

[31] Deng Y, Liu W, Liu K, et al. Clinical characteristics of fatal and recovered cases of coronavirus disease 2019 (COVID-19) in Wuhan, China: a retrospective study [published online ahead of print, 2020 Mar 20]. Chin Med J (Engl) [Internet]. 2020;10.1097/ CM9. 0000000 000000824.

[32] Zhou F, Yu T, Du R, et al. Clinical course and risk factors for mortality of adult inpatients with COVID-19 in Wuhan, China: a retrospective cohort study. Lancet [Internet]. 2020; 395(10229):1054–1062. doi:10.1016/S0140-6736(20)30566-3.

[33] Fine MJ, Auble TE, Yealy DM, et al. A prediction rule to identify low-risk patients with community-acquired pneumonia. N Engl J Med 1997;336(4):243–50.

[34] Bone RC, Sibbald WJ, Sprung CL. The ACCP-SCCM consensus conference on sepsis and organ failure. Chest 1992;101(6):1481–3.

[35] Guan WJ, Ni ZY, Hu Y, et al. China medical treatment expert group for COVID-19. Clinical characteristics of coronavirus disease 2019 in China. N Engl J Med 2020; 382: 1708–1720. https://doi.org/10.1056/ NEJMoa2002032 [Epub ahead of print].

[36] Wang D, Hu B, Hu C, et al. Clinical characteristics of 138 hospitalized patients with 2019 novel coronavirus-infected pneumonia in Wuhan, China. JAMA 2020 (Feb 7); 323: 1061. https://doi.org/10.1001/jama. 2020.1585. [Epub ahead of print].

[37] Myers LC, Parodi SM, Escobar GJ, et al Characteristics of hospitalized adults with COVID-19 in an integrated health care system in California. JAMA (Apr 24) 2020;e207202. https://doi.org/10.1001/jama. 2020.7202.

